# Shielding individuals at high risk of COVID-19: a micro-simulation study

**DOI:** 10.1101/2022.01.03.22268675

**Authors:** Kevin van Zandvoort, Caroline Favas, Francesco Checchi

## Abstract

**Background:** One of the proposed interventions for mitigating COVID-19 epidemics, particularly in low-income and crisis-affected settings, is to physically isolate individuals known to be at high risk of severe disease and death due to age or co-morbidities. This intervention, known as ‘shielding’, could be implemented in various ways. If shielded people are grouped together in residences and isolation is imperfect, any introduction of infections within the shielding group could cause substantial mortality and thus negate the intervention’s benefits. We explored the effectiveness of shielding under various modalities of implementation and considered mitigation measures to reduce its possible harms.

**Methods:** We used an individual-based mathematical model to simulate the evolution of a COVID-19 epidemic in a population of which a fraction above a given age cut-off are relocated to shielding residences, in which they have variable levels of contacts with their original household, the outside world and fellow shielding residents. We set our simulation with the context of an internally displaced persons’ camp in Somaliland, for which we had recently collected data on household demographics and social mixing patterns. We compared an unmitigated epidemic with a shielding intervention accompanied by various measures to reduce the risk of virus introduction and spread within the shielding residences. We did sensitivity analyses to explore parameters such as residence size, reduction in contacts, basic reproduction number, and prior immunity in the population.

**Results:** Shielded residences are likely to be breached with infection during the outbreak. Nonetheless, shielding can be effective in preventing COVID-19 infections in the shielded population. The effectiveness of shielding is mostly affected by the size of the shielded residence, and by the degree by which contacts between shielded and unshielded individuals are reduced. Reductions in contacts between shielded individuals could further increase the effectiveness of shielding, but is only effective in larger shielded residences. Large shielded residences increase the risk of infection, unless very large reductions in contacts can be achieved. In epidemics with a lower reproduction number, the effectiveness of shielding could be negative effectiveness.

**Discussion:** Shielding could be an effective method to protect the most at-risk individuals. It should be considered where other measures cannot easily be implemented, but with attention to the epidemiological situation. Shielding should only be implemented through small to medium-sized shielding residences, with appropriate mitigation measures such as reduced contact intensity between shielded individuals and self-isolation of cases to prevent subsequent spread.

## Introduction

COVID-19 epidemics may prove particularly difficult to manage in low-income and crisis-affected settings of the world, where resources to scale up case management are insufficient to meet demand, insufficient public health and laboratory capacity precludes effective use of testing and contact tracing, and socio-economic circumstances (e.g. overcrowding, inadequate water and sanitation, imperative to generate an income) make it difficult for populations to adopt and sustain physical distancing^1,2^. Over the next years, waning immunity, novel variants, and insufficient vaccine access could expose populations to renewed COVID-19 waves. This suggests a need to identify options for mitigating epidemics affecting these populations that do not require socially and economically harmful lockdown measures. One such option, known as ‘shielding’, consists of physically isolating individuals known to be at high risk of developing severe disease and dying if infected with SARS-CoV-2^2^. This intervention could reduce severe disease and mortality while herd immunity builds up in the low-risk population groups^3^; it would also lessen health service pressure and enable societies and economies to remain functional.

While shielding could best be viewed as a community-led and -designed intervention with no pre-set modalities, we have previously suggested that likely arrangements could include grouping high-risk individuals together into ‘shielding residences’, particularly where individual shielding within households is impracticable^4^. The number of people shielded together, as well as various other characteristics of the intervention (e.g. its timing of introduction; arrangements for infection prevention and control) could also vary.

While compartmental dynamic models indicate that shielding has a substantial potential to reduce mortality and health service pressure^3^, such models do not fully capture the individual-level dynamics of the intervention: in particular, what remains unexplored is the potential harm of inadvertently introducing infection into shielded residences. This harm might or might not be outweighed by the benefit of shielding, compared to no mitigation, and could itself be mitigated through a number of measures, such as not grouping too many high-risk people together; only shielding people who are not symptomatic; and supporting people to isolate if they develop symptoms while in a shielding residence.

In order to support the design of appropriate, safe shielding interventions, we used mathematical modelling to generate quantitative predictions of the potential benefit and harms of shielding people together under different scenarios of density of shielded residents, timing of shielding implementation with respect to local epidemic onset, compliance with physical isolation and interventions to mitigate the risk of outbreaks within shielding residences. Harm here is defined as infection: it is assumed that high-risk individuals, once infected, would have a high probability of severe disease and death, as described elsewhere^3,5^.

## Methods

### Model structure

We used an individual-based, probabilistic mathematical model (IBM) to study the research question. We set our model within the real-life setting of Digaale internally displaced persons’ (IDP) camp near Hargeisa, Somaliland, for which data on demographics and other relevant parameters was recently collected.^6^ While most IBMs attempt to answer population-level questions (e.g. the potential impact of an intervention), they do so by simulating the trajectories of each individual across different infection and disease states, as a function of various individual-level characteristics. They are particularly useful to simulate small population units (e.g. households) in which chance is very influential.

In the model, individuals fall within *a* age groups with relative proportions *p*_*i*_ (where *i* is a given age group). Individuals live in any of *H* households of mean size *m*_*h*_ (we use the empirically measured *H* and household sizes *N*_*h*_). The population of size *N* is seeded with a single SARS-CoV-2 infection at the start of the simulation. The IBM is ran for a total of 365 days or until the outbreak naturally dies out.

Given its short timeframe, births, deaths, ageing and migration are omitted from the model. The endpoint of interest is infection (i.e. the effectiveness of shielding relates to its ability to reduce infection risk among high-risk individuals): as such, the clinical outcome and treatment status of cases are not included in this model.

### States and transitions

At any time *t*, individualsare within one of the following epidemiological states: *S* (susceptible), *E* (pre-infectious), *I*_*P*_ (infectious but pre-symptomatic), *I*_*C*_ (infectious and symptomatic), *I*_*S*_ (infectious and asymptomatic throughout the infection), or *R* (removed: recovered and assumed to be immune or deceased). The age-specific probability of becoming a symptomatic case is *y*_*i*_.

Over any *Δt* time unit, any given individual *x* has the following binomial probabilities of transitioning to a subsequent state:

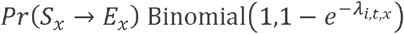

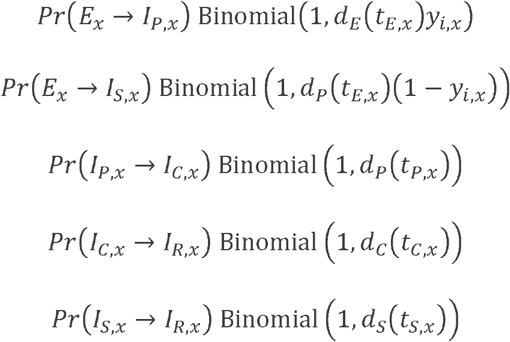

where 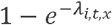 is the age-specific instantaneous force of infection (expressed as an incidence risk) experienced by a susceptible individual, as detailed below; and *d*_*E*_, *d*_*P*_, *d*_*C*_ and *d*_*S*_ are cumulative distribution functions (CDFs) for the duration of the corresponding states: *d*_*E*_*(t*_*E,X*_) denotes the CDF for the duration of the pre-infectious state evaluated at the time already spent by individual *x* in that state, and so on.

### Transmission dynamics

Over any *Δt* time unit, individuals *x* of any age *i* within each household *h* move from *S* to *E* based on an individual-specific instantaneous force of infection that is the sum of *λ* due to contacts within the household and *λ* due to extra-household contacts:

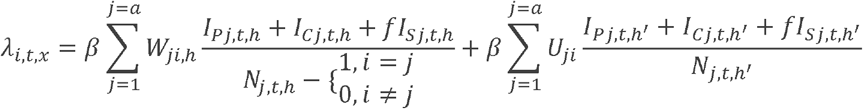

where *β* is the probability of infection per infectious contact, *f* is the relative infectiousness of asymptomatic infections, compared to cases that do develop symptoms, *W* is the matrix of per-capita contact rates within household *h* among individuals of ages *i* and *j*, and *U* is the corresponding contact matrix outside the household (*h*′ denotes individuals in the population excluding the household itself). *W* + *U = Z*, i.e. the full contact matrix. We assume random mixing of individuals within households, and the intra-household term simplifies to

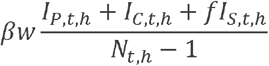

where 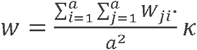 is the mean per-capita intra-household contact rate, and *κ* is the ratio between the dominant eigenvalues of the reported intra-household contact matrix and the expected intra-household contact matrix, the latter being based on all household age pairs in the modelled population assuming each household age pair makes one contact per day.

The basic reproduction number *R*_*0*_ is defined as the average number of secondary infections generated by a typical infected individual in a fully susceptible population and may be computed as the absolute value of the dominant eigenvalue of the next generation matrix (NGM) of the corresponding compartmental model structure, defined as

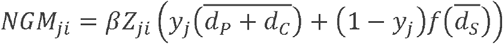

where accents indicate the expected (average) values. Lastly, *β* is the ratio of this eigenvalue and the *R*_*0*_ value assumed in the simulation (see below). *R*_*0*_ values were validated by running multiple model iterations with different seeds to start the random number generator, and by calculating the average number of secondary cases derived from all infectious individuals who completed their period of infectiousness in the first 30 days of the simulation.

Model simulations start at day 1, when one randomly selected individual living in the camp is made infectious.

### Shielding and related interventions

Shielding is introduced when the prevalence of symptomatic cases in the population reaches 5 per 1000 individuals, by relocating a proportion *p*_*g*_ of high-risk individuals within *H* (for simplicity, high risk is defined based solely on an age cut-off, i.e. ≥ 60 years old) into *G* shielded residences containing a variable maximum number *N*_*g*_ of high-risk residents.

While shielded, high-risk residents’ contact with other people remains structured as above, but is reduced or increased to varying extents as follows:

- contact with unshielded members of the household of origin is reduced by a factor *σ*_*w*_;
- contact with other unshielded people is reduced by a factor *σ*_*u*_;
- contact with other shielded individuals within *g* is assumed to occur by random mixing, with *w* as the baseline per-capita rate, either reduced (if physical distancing and improved hygiene are also maintained within *g*) or increased (if, vice versa, *g* is overcrowded and unsanitary) by a factor *θ*_*w*_.

Accordingly, each shielded individual originally from household *h* experiences an age-specific instantaneous force of infection

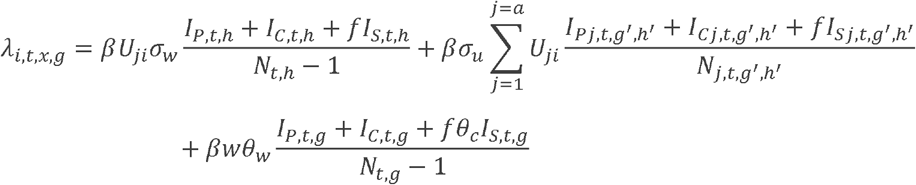

where *g*′ denotes individuals not living within the shielded residence. We assume no transmission between different shielded residences and random mixing among shielded individuals (in practice, it is plausible that people might mix preferentially with close family or acquaintances or those whom they room most proximately to).

Outside *g*, all unshielded individuals experience the following force of infection:

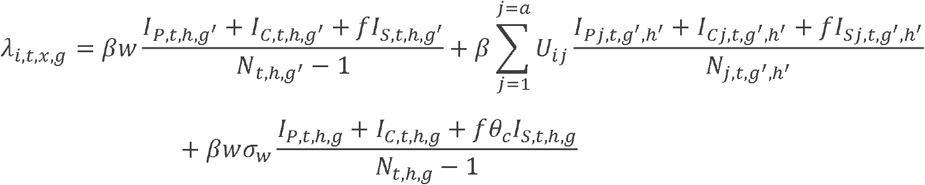

We assume that unshielded individuals only make effective contact with shielded residents from their own household.

If shielded residents are infected (any *E* or *I* class) at the time of shielding, or become infected while shielded, they may infect other residents. The following measures to mitigate the occurrence and size of such outbreaks are contemplated in the model:

1. Nothing is done;
2. Varying extent of isolation from people outside *g*, resulting in values of *σ*_*w*_ and *σ*_*u*_ tending towards 0; for simplicity, we assume *σ*_*w*_ = *σ*_*u*_ = *σ*;
3. Physical distancing and hygiene improvements within *g*, resulting in a value of *θ*_*w*_ < 1;
4. Symptomatic cases within *g* self-isolate, causing a further reduction *θ*_*c*_ in all their contacts;
5. Symptomatic cases within *g* exit the shielded residence with a delay *δ* since symptom onset, and return to their households (we assume this is a more likely prospect than hospitalisation in most low-income or crisis-affected settings; either way, the destination on exit has a negligible effect on the model);
6. As soon as a symptomatic case occurs within *g*, all residents exit shielding and return to their households;
7. At the time shielding is introduced, any high-risk individuals who are symptomatic or who live in a household with at least one symptomatic case remain in their households and only join *g* once they and all their household members are recovered.
8. At the time shielding is introduced, all high-risk individuals are tested irrespective of symptoms, and any positive cases do not enter the shielding accommodation, with a test sensitivity *ρ*.
9. Shielding is not implemented at all.

### Parameter values and data sources

Table 1 lists model parameters and their input values.

**Table 1.** Model parameters and their values or ranges.

| Parameter | Description | Value (base scenario in bold) | Source |
| --- | --- | --- | --- |
| $H$ | Number of households | **715** | <sup>6</sup> |
| $i, j$ | Age strata in years (number of age strata = $a$) | 0-9, 10-19, 20-29, 30-39, 40-49, 50-59, 60-69, 70-79, 80+ | <sup>6</sup> |
| $p_i$ | Proportion of people in each age stratum $i$ | Resampled within each model iteration | <sup>6</sup> |
| $m_h$ | Mean household size | Resampled within each model iteration | <sup>6</sup> |
| $N_h$ | Number of people in each household $h$ | Resampled within each model iteration | <sup>6</sup> |
| $N_g$ | Number of people living together within each shielded residence | Variable: 2, 4, 8, 16 | n/a |
| $\Delta t$ | Time step for discrete-time simulation | 1 day | n/a |
| $d_E$ | Latent period in days | $\sim \text{gamma}(\mu = 2.5, k = 4)$ | <sup>7-9</sup> |
| $d_P$ | Duration of pre-symptomatic infectiousness in days | $\sim \text{gamma}(\mu = 2.5, k = 4)$ | <sup>10</sup> |
| $d_C$ | Duration of symptomatic infectiousness in days | $\sim \text{gamma}(\mu = 2.5, k = 4)$ | <sup>7-9</sup> |
| $d_S$ | Duration of asymptomatic infectiousness in days | $\sim \text{gamma}(\mu = 5, k = 4)$ | Assumed to be the same as duration of total infectious period for clinical cases |
| $y_i$ | Probability of becoming a symptomatic case, if infected, for age group $i$ | Age-dependent, as estimated in Davies et al. | <sup>11</sup> |
| $R_0$ | Basic reproduction number | Variable: 1.5, **2.5**, 3.5, 5 | Assumption, based on a global distribution of $R_0$ values for the initial SARS-CoV-2 virus<sup>12</sup> and mean estimate for the Delta variant<sup>13</sup> |
| $\Psi$ | Proportion immune at the start of the simulation | Variable: 0, 25%, 50% | Assumption |
| $f$ | Relative infectiousness of asymptomatic cases | 50% | Assumption |
| $\omega$ | Within-household per-capita contact rate | Ratio between dominant eigenvalue of expected within household contact matrix and reported within household contact matrix | <sup>6</sup> |
| $U$ | Age-dependent contact matrix outside the household | Resampled within each model iteration | <sup>6</sup> |
| $Z$ | Age-dependent contact matrix within and outside of the household | Resampled within each model iteration | <sup>6</sup> |
| $\beta$ | Probability of transmission per contact with an infectious individual | See text | Computed within the model |
| $p_g$ | Proportion of people eligible for shielding who are shielded | Variable: 0.0, 0.2, 0.4, 0.6, **0.8**, 1.0 | n/a |
| $\sigma$ | Contact between shielded and unshielded individuals, relative to pre-shielding | Variable: 0.0, **0.2**, 0.4, 0.6, 0.8, 1.0 | n/a |
| $\theta_w$ | Change in contact intensity among shielded individuals, relative to within-household contacts | Variable: 0, 0.2, 0.4, 0.6, 0.8, **1.0**, 1.2 | n/a |
| $\theta_c$ | Contact between symptomatic shielded individuals and other shielded individuals, relative to before becoming symptomatic | Variable: 0, 0.2, 0.4, 0.6, 0.8, **1.0** | n/a |
| $\delta$ | Delay in days between onset of symptoms and exiting the shielded residence | Variable: **N/A**, 0, 2, 4 | n/a |
| $\rho_E$ | Sensitivity of SARS-CoV-2 laboratory test among individuals in pre-infectious class | 0 | assumption |
| $\rho_P$ | Sensitivity of SARS-CoV-2 2 lateral flow test among individuals in pre-symptomatic infectious class | 58.7% | <sup>14</sup> |
| $\rho_C$ | Sensitivity of SARS-CoV-2 lateral flow test among individuals in symptomatic infectious class | 84.2% | <sup>14</sup> |
| $\rho_S$ | Sensitivity of SARS-CoV-2 2 lateral flow test among individuals in asymptomatic infectious class | 58.7% | <sup>14</sup> |

Data on social contacts were taken from a cross-sectional survey conducted among 501 individuals living in Digaale IDP camp, Somaliland.^6^ The IDP camp is a permanent settlement established in 2014, has an area of approximately 150 hectares, and is situated 4km from Hargeisa, the capital city. There are an estimated 715 inhabited shelters in Digaale, with an average household size of 4.4 individuals. The median age of residents is 15 years (interquartile range 7 to 34). 7.5% of the population is aged 60 years or older.

We simulated a total of 715 households using the Digaale data, where we randomly sampled households with replacement from the set of all households and household members in the empirical data. The population structure and contact matrix for non-household contacts was resampled in every iteration of the model. To ensure comparability between scenarios, the same population structure and random seed were used for all modelled scenarios within the same iteration of the model.

### Analysis

We implemented 1000 iterations of the model to *T* = 12 months for four hypothetical scenarios of *N*_*g*_ (2, 4, 8, 16), and tracked the following outcomes for each scenario:

- Proportion of shielded residences that are breached, i.e. where at least one person becomes infected after moving into the shielded residence, by shielding mitigation measure (1-8 above);
- Cumulative risk of infection among high-risk individuals over *T*, including unshielded and shielded person-time, by mitigation measure (1-8) and under no shielding (9);
- Percent reduction in cumulative risk of infection among high-risk individuals within *G*, by mitigation measure (2-8), compared to no shielding mitigation (1);

We only included model iterations where a minimum outbreak size of 50 infections was reached in the corresponding unmitigated scenario.

The median and 95% percentile interval of all simulations are presented. Scenarios are directly compared against scenarios within the same model iteration.

We assumed an *R*_0_ value of 2.5 with no prior immunity for our baseline scenario. To assess the sensitivity of our results to new variants with high transmissibility, or pre-existing immunity, we ran sensitivity analyses for *R*_0_ and Ψ, the proportion of the population already immune to infection at the start of the simulation. We also ran sensitivity analysis for *p*_*g*_, the proportion of eligible high-risk people who are shielded.

## Results

### Risk of virus introduction into shielded residences

Generally, we project a high probability that shielding residences are breached during the outbreak, i.e. at least one resident is newly infected when shielded (Figure 1). This risk increases proportionally to the shielded residence size. Assuming an 80% reduction in contacts between shielded and unshielded individuals, there would be a 38% (29 - 49) risk of any shielded residence of size two being breached, whereas this risk increases proportionally to the shielded residence size. Breaching risk is moderated by the effectiveness of shielding: the extent to which contacts between shielded and unshielded contacts are reduced. Very effective implementations of shielding, where contacts with unshielded individuals are reduced by 100% and onward transmission can only result from pre-shielding infections, protect nearly all small shielded residences, but cannot prevent infections to occur in larger shielded residences.

**Figure 1.**
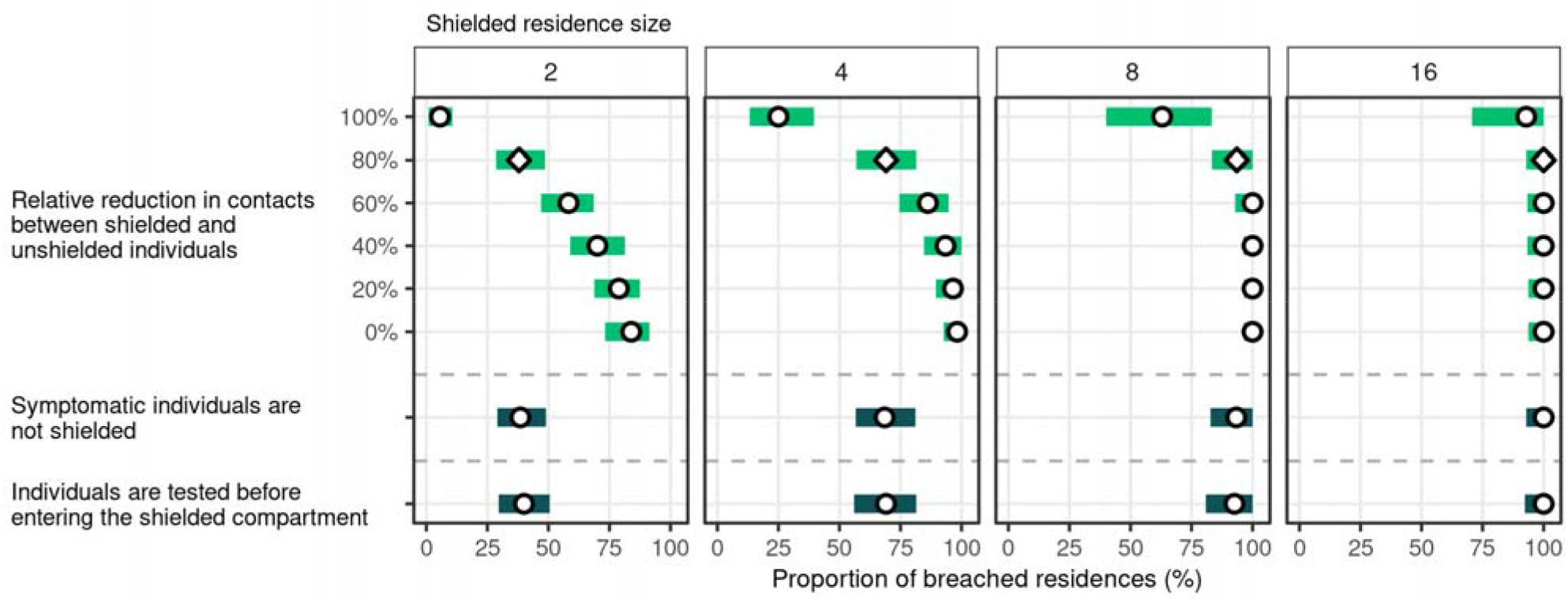
Cumulative risk of breaching the shield over the entire period of the outbreak, by shielded residence size (in facet columns) and mitigation measure (y-axis). Points represent median estimates over 1000 model runs, while bars represent their 95% uncertainty intervals. In the baseline scenario (diamond), contacts between shielded and unshielded individuals are decreased by 80% and no other mitigation measures are implemented. Estimates for the two mitigation measures assume the same reduction in contacts as the baseline scenario, in addition to the implemented mitigation measure. In all scenarios shown, *R*_0_ was 2.5 and 0% of the population were immune at the start of the simulation.

Additional mitigation measures that aim to screen individuals for infection before they were shielded were not effective in reducing the risk that shielded residences are shielded. In none of the scenarios considered was the proportion of breached shielded residences significantly different when either syndromic screening or testing was implemented prior to screening. There was only one scenario (with *R*_0_ of 1.5, 25% prior immunity, 20% of the high-risk population that was shielded, and a shielded residence size of 2) where testing but not syndromic screening significantly increased the mean number of days until shielded residences were shielded by 3 days (0 - 12).

### Effectiveness of shielding against individual risk of infection

High-risk individuals who are not shielded experience similar infection rates as unshielded low-risk individuals (Figure 2) over time. High-risk individuals who are shielded can have substantially lower risks of infection, depending on the implementation of shielding.

**Figure 2.**
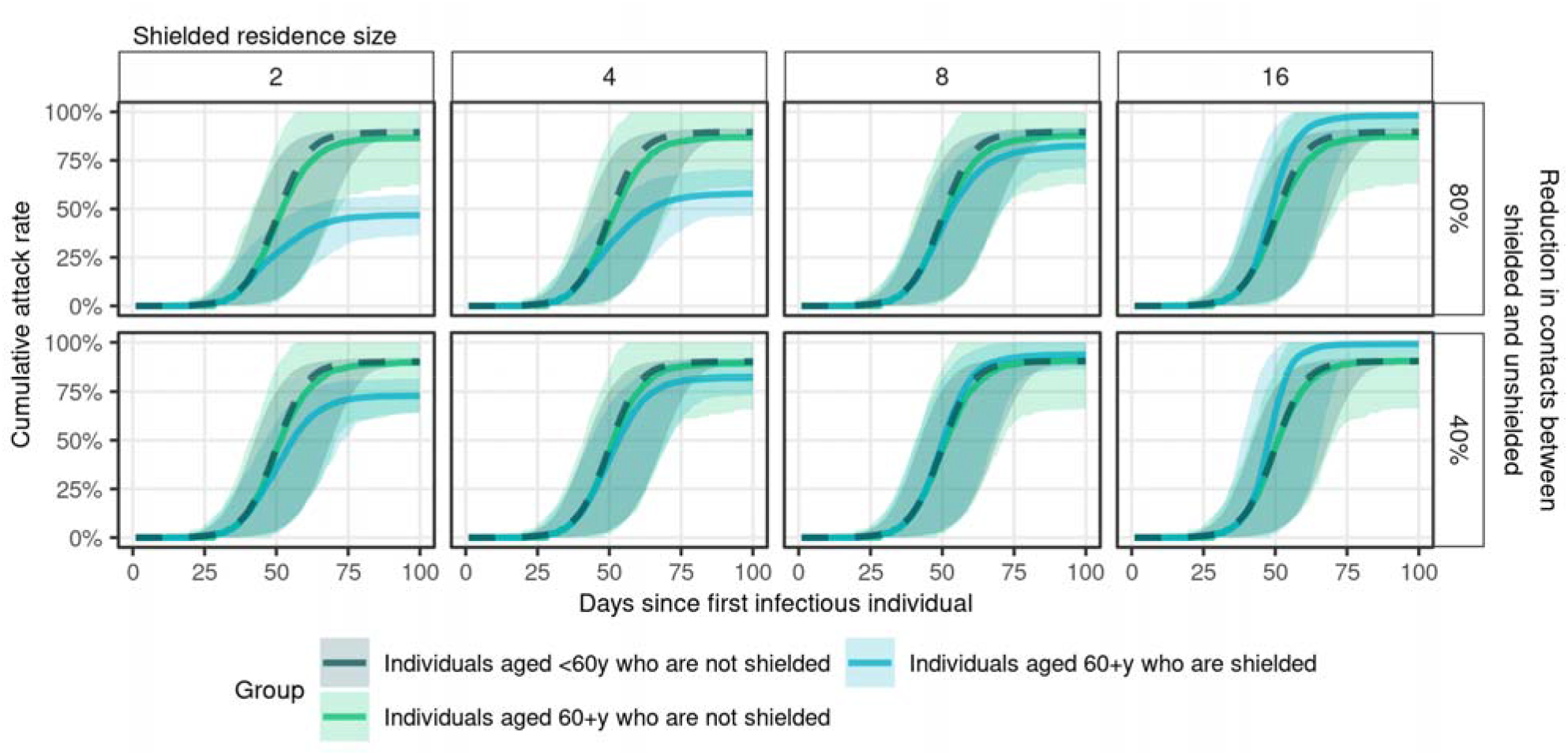
Cumulative infection risk by shielded status over time, since SARS-CoV-2 introduction in the IDP camp. Estimates are stratified by shielded residence size (in facet columns) and reduction in contacts between shielded and unshielded individuals (in facet rows). In each scenario, lines represent the median attack rate across 1000 model runs at each timepoint, whereas corresponding 95% uncertainty intervals are shown by shaded areas of the same colour. Estimates for (unshielded) low-risk individuals are shown in dark-green and a dashed line. Unshielded high-risk individuals are shown in light-green, while shielded high-risk individuals are shown in blue. In all scenarios shown, *R*_0_ was 2.5, 0% of the population were immune at the start of the simulation, and 80% of the high-risk population was shielded.

The risk of infection for shielded individuals increases with the size of the shielded residences, and is mitigated by the degree by which contacts between shielded and unshielded individuals are reduced. Shielding has no substantial effect on infection risk of unshielded individuals, regardless of the proportion of high-risk individuals that is shielded (Supplemental Figure 2). Shielding does not substantially alter the timing of outbreaks within the shielded population, and has no substantial impact on infections in the unshielded low-risk population. Individuals who are shielded in large shielded residences could experience a higher risk of infection compared to unshielded individuals of the same age, even with relatively effective implementations of shielding.

Figure 3 shows the effectiveness of shielding to prevent infections in high-risk individuals when compared to unmitigated epidemics for a range of different shielding implementations. Effectiveness is calculated as the relative reduction in the cumulative number of infections during the entire modelled outbreak. Even if shielding would not be effective in reducing contacts with unshielded individuals at all, rehousing individuals in shielded residences of size 2 (which is half the median household size) would result in a 10% (3 to 17) reduction of cases. As the size of the shielded residence increases, higher reductions in contacts between shielded and unshielded individuals are needed to achieve substantial reductions in infection risk, whereas even a 100% reduction in contacts is insufficient to reach any substantial reduction (2%; -9 to 25) in the absence of other mitigation measures for shielded residences of size 16. For less effective implementations of shielding, rehousing high-risk individuals in residences of size 16 may even increase their risk of infection: the effectiveness of shielding is -6% (−13 to -2) when contacts between shielded and unshielded individuals are reduced by 60%.

**Figure 3.**
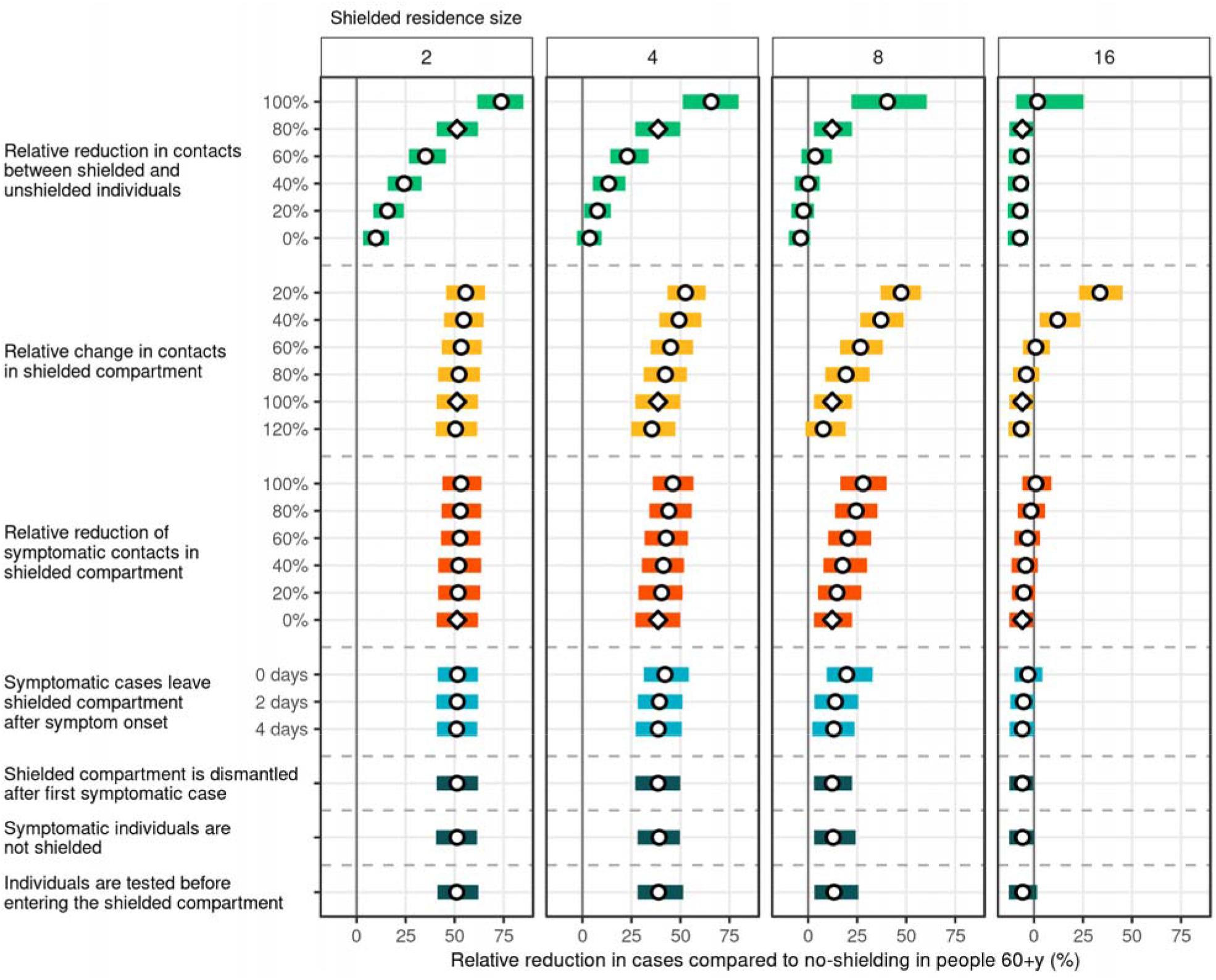
Relative reduction in infections in shielded high-risk individuals compared to no-shielding, by shielded residence size (in facet columns) and shielded scenario (y-axis). Points represent median estimates over 1000 model runs, while bars represent their corresponding 95% uncertainty intervals. In the baseline scenario (diamonds), contacts between shielded and unshielded individuals are decreased by 80%, contacts between shielded individuals within the same shielded residence remain unchanged, symptomatic individuals who are shielded do not reduce their effective contacts, and no other mitigation measures are implemented. In all other scenarios (circles), only a single parameter is changed with respect to the baseline scenario. Estimates to the left of the thick vertical grey line correspond to negative effectiveness: an increase in infections after shielding compared to no shielding. In all scenarios, *R*_0_ was 2.5, 0% of the population were immune at the start of the simulation, and 100% of high-risk individuals were shielded.

As the size of the shielded residence increases, the relative change in contacts between shielded individuals living in the same residence, relative to the average contact intensity between individuals within a household, becomes more important for infection control. In shielded residences of size 8, reducing contacts within the shielded compartment by 40% (in addition to the 80% reduction of contacts with unshielded individuals) may increase the effectiveness of shielding from 12% (3 to 22) to 27% (16 to 38). For a residence of size 16, a reduction of at least 60% would be required in order for shielding to have a beneficial effect of 12% (3 to 24). Contact intensity between shielded individuals only marginally affects the risk when individuals are shielded in smaller residences (2 to 4 individuals). If relative contact intensity between shielded individuals were to increase rather than decrease, the benefit of shielding in shielding residences of moderate sizes decreases, though this extent is mitigated by residence size.

Self-isolation of symptomatic cases who are shielded could further increase the effectiveness of shielding, though it is less effective than an overall reduction in contact intensity within the shielded residence. The largest effects are present in shielded residences of size 8, whereas effects are small for smaller and larger residences. The impact of other mitigation on the effectiveness of shielding is small. If symptomatic cases would leave the shielded residence on the first day they develop symptoms, effectiveness increases from 39% (27 to 50) under the baseline strategy to 42% (31 to 54) when shielded residences are of size 4, and from 12% (3 to 22) to 20% (9 to 33) when shielded residences are of size 8, but any benefit disappears if there is a delay of at least two days. Testing for infection or syndromic screening prior to shielding is not effective in reducing infection risk in the shielded population.

### Sensitivity to *R*_0_ and pre-existing immunity

Our main analysis has assumed an *R*_0_ of 2.5, but we assessed the sensitivity of this assumption to our results in scenarios where *R*_0_ was 1.5, 3.5, or 5. In addition, whereas our main analysis assumed that all individuals in the population were susceptible, we assessed the sensitivity of our results to scenarios where 25% or 50% of the population was immune to SARS-CoV-2 infection. Table 2 lists the median attack rates in unmitigated outbreaks under each scenario.

**Table 2.** Infection attack rates in high-risk individuals, by *R*_0_ and prior immunity.

| | $R_0$ | | | |
| --- | --- | --- | --- | --- |
| Prior immunity | 1.5 | 2.5 | 3.5 | 5 |
| 0% | 72% (63 - 80) | 93% (88 - 96) | 98% (95 - 100) | 100% (98 - 100) |
| 25% | 29% (4 - 41) | 59% (51 - 67) | 68% (61 - 74) | 72% (66 - 79) |
| 50% | 2% (1 - 4) | 18% (2 - 27) | 33% (25 - 41) | 41% (34 - 49) |

Supplemental Figure 1 shows the proportion of breached shielded residences by shielded residence size, *R*_0_, and reduction in contacts between shielded and unshielded individuals. Under all levels of *R*_0_ considered, there is a high risk that shielded residences of size 16 would be breached, with 71% (46 - 93) of residences breached when contacts with unshielded individuals are reduced by 100% and *R*_0_ is only 1.5. Only residences of size 2 result in a low number of breached residences in all scenarios. High effectiveness of shielding (≥80% reduction in contacts with unshielded individuals) and small shielded residences (2 to 4 people) are needed to prevent a substantial number of residences being breached for outbreaks with a high *R*_0_ (≥ 3.5).

Regardless of the *R*_0_, unshielded low- and high-risk individuals experience similar attack rates over time (Supplemental Figure 2). A large shielded residence results in high cumulative attack rates in all scenarios considered. In scenarios with a large shielded residence size (16) and low *R*_0_, the cumulative attack rate is substantially higher in shielded compared to unshielded individuals. The cumulative attack rates in all groups is not affected by the proportion of high-risk individuals that are shielded.

A shielded residence size of 2 did never result in a negative impact of shielding when compared to unmitigated scenarios in any of the scenarios considered. Scenarios with low effective reproductive numbers, either through a low initial *R*_0_ or a higher *R*_0_ with a substantial proportion of the population already immune, could result in a negative impact of shielding. Without any pre-existing immunity in the population and an *R*_0_ of 1.5, a shielded residence size of 16 would result in a negative impact in all scenarios where the reduction in contacts between shielded and unshielded individuals was at most 60%.

While the maximum effectiveness of shielding compared to unmitigated scenarios decreases as the proportion of the population that is already immune increases, more leaky implementations of shielding (a lower reduction in contacts between shielded and unshielded individuals) increase in their effectiveness as the proportion that is already immune increases. For instance, when *R*_0_ is 5, shielding in residences of size 2 with 100% reduction in contacts between shielded and unshielded individuals prevents 60% (45 - 75) of cases with 0% pre-existing immunity, 60% (44 - 74) with 25% pre-existing immunity, and 46% (25 - 66) with 50% pre-existing immunity. Shielding in residences of size 2 with only 40% reduction in contacts prevents 7% (3 - 12) of cases with 0% pre-existing immunity, 14% (7 - 23) with 25% pre-existing immunity, and 21% (8 - 33) with 50% pre-existing immunity.

## Discussion

### Main findings

The potentially devastating impact of COVID-19 on humanitarian settings has been highlighted many times^15–18^, but few empirical and modelling studies have focussed on these settings due to a lack of available data^19^. To our knowledge, our study is the first modelling study to specifically look at shielding as a COVID-19 mitigation option for IDPs or camp-like settings. Our results could be extended to other low resource settings with low vaccine coverage, where large scaled lockdowns cannot be sustained long term.

We found that shielding can be an effective measure to protect individuals at high risk of morbidity and mortality due to COVID-19 using demographic and contact data from Digaale IDP camp, though there is a risk of harm in specific scenarios. The impact of shielding largely depends on how effectively it is implemented, requiring stringent isolation of shielded people, and high uptake (coverage) for substantial population impact.

Specifically, the effectiveness of shielding mostly depends on two factors, i. the total number of individuals who are shielded together, and ii. the reduction in contact between shielded and unshielded individuals. This is not surprising, as the effectiveness of public health and social measures largely depends on changing existing social contact structures, which are impacted by these two factors.

Generally, our model projects that smaller shielding residences would be considerably more effective than larger ones. They are less likely to be breached, and there is only limited opportunity for onward transmission when breached. These small-scale implementations of shielding may also be more acceptable to implement: shielding was seen as an acceptable method to protect the most vulnerable in Sudan, though extra-household shielding arrangements were generally viewed to be socially unacceptable^20^. Shielding within or close to the old household may result in more intense care by low-risk individuals to occur, where contact rates with unshielded individuals in general cannot be reduced by high levels.

Large shielded residences should be avoided, as they were not effective in all scenarios unless a very stringent form of shielding is implemented, with none or very little contact between shielded and unshielded individuals and among shielded individuals within the same shielding residence. This would be a potentially daunting standard to achieve in real-life application, as has been shown in the control of COVID-19 in nursing homes.^21^

In settings where it is not feasible to implement small-group shielding strategies, shielding in medium sized shielding residences could be an alternative. However, shielding in medium sized shielding residences would highly benefit from further mitigation measures, such as a reduction in contact intensity between shielded individuals living in the same shielding residence, and self-isolation or quarantine of shielded cases once any resident becomes symptomatic. Contact intensity may well be lower when individuals from different households are housed together, compared to when individuals who are shielded together only belong from the same household.

If self-isolation in a given shielding residence is unfeasible, syndromic screening may be an effective alternative where symptomatic individuals exit the shielding residence as soon as possible after symptom onset. However, as COVID-19 symptoms are highly non-specific^22^, this would ideally be combined with testing strategies, and symptomatic individuals would ideally be quarantined.

Our model predicts little effect of other mitigation measures considered, such as testing individuals before they are shielded, or dismantling the shielding residence once a case arises. Both measures are most likely to detect current infections, but would miss any individuals already infected but not yet infectious. Continuous testing may be an effective alternative^23^, but was not considered in our analysis, as (asymptomatic) testing capacity may be limited in humanitarian settings.

When *R* is low, either as a result of a low initial *R*_0_, or pre-existing immunity in the population through natural immunity or vaccination, shielding individuals in medium- to large groups could increase their infection risk and result in harm. The epidemiological situation should be considered to assess the appropriateness of shielding in any setting.

### Limitations

There are several limitations to this study. Although we were able to use baseline empirical contact-data, these contact patterns could be further altered if shielding would be implemented and contact dynamics change.

We only considered Digaale IDP camp, as to our knowledge it is the only humanitarian setting for which detailed contact and demographic data is available. Social contacts are context specific^24^, and Digaale is a relatively small-scale peri-urban settlement, which may not be representative of other low-resource or crisis-affected settings. Our model did not include an additional force of infection from outside the camp, beyond the first seeding event. Fewer than 2% of all contacts were reported to be made outside of the camp in the contact data, so the relative impact of additional transmission from outside the camp in the unshielded population is expected to be small once transmission has already been established.

We did not estimate the number of cases and deaths that would be expected after infection, though these are proportional to the number of infections. We focussed our analysis on the impact of shielding high-risk individuals, but only assumed individuals aged 60+ years old to be at high-risk. Although the age-risk profile may well be different in low-resource settings compared to stable settings^25^, empirical estimates are missing, and similar levels of effectiveness could be expected with broader definitions of high-risk groups.

We did not consider simultaneous implementations of other measures, such as physical distancing in the general population. In reality, shielding would most likely be implemented in addition to other non-pharmaceutical interventions. We previously found that a combination of shielding, self-isolation, and moderate social distancing may be an effective and feasible strategy for low-income countries^3^. Whereas large-scaled lockdowns could temporarily be effective in delaying epidemic peaks^3,26^, these cannot sustainably be implemented in most humanitarian responses and low-income countries.

## Conclusions and recommendations

COVID-19 epidemics could be difficult to control in low-resource settings, including humanitarian settings, where residents live in crowded spaces, little healthcare capacity is available, and stringent social distancing measures cannot sustainably be implemented. Shielding could be an effective alternative to protect high-risk individuals, when it is not possible to implement more stringent factors such as lockdowns.

Where possible, shielding should be implemented through small to medium-sized shielding residences, with appropriate mitigation measures such as reduced contact intensity between shielded individuals and self-isolation of cases to prevent subsequent spread. Shielding should not be implemented if only large-sized shielding residences are available, as this could result in an increased infection risk for high-risk individuals and do more harm than good.

## Data Availability

All code used is available online at https://github.com/kevinvzandvoort/covid19-shielding-microsimulation-digaale

## Supplementary material

**Supplemental Figure 1.**
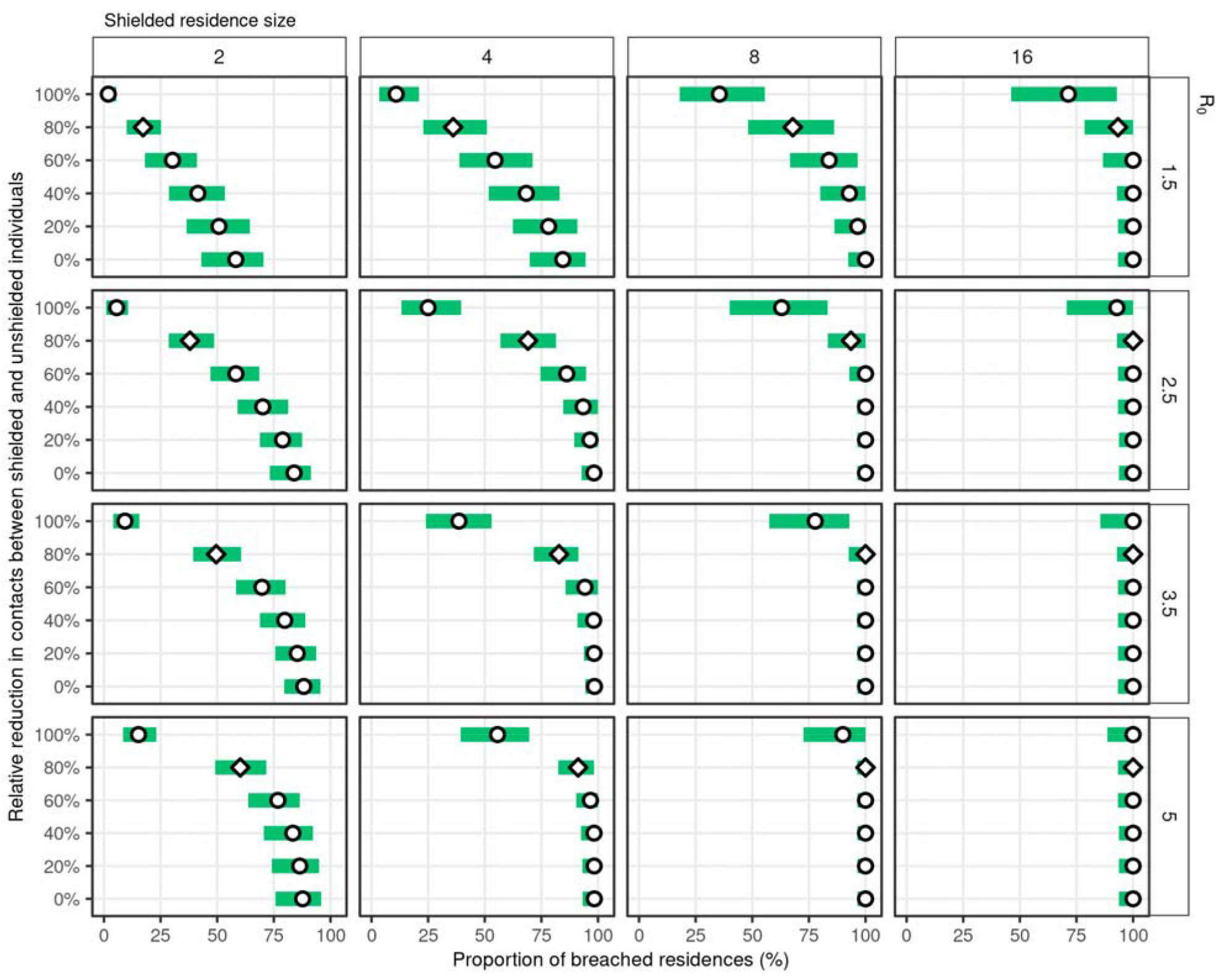
Cumulative risk of breaching the shield, by shielded residence size (in facet columns) and *R*_0_ (y-axis). Points represent median estimates over 1000 model runs, while bars represent their 95% uncertainty intervals. In all scenarios shown, 0% of the population were immune at the start of the simulation.

**Figure 2.**
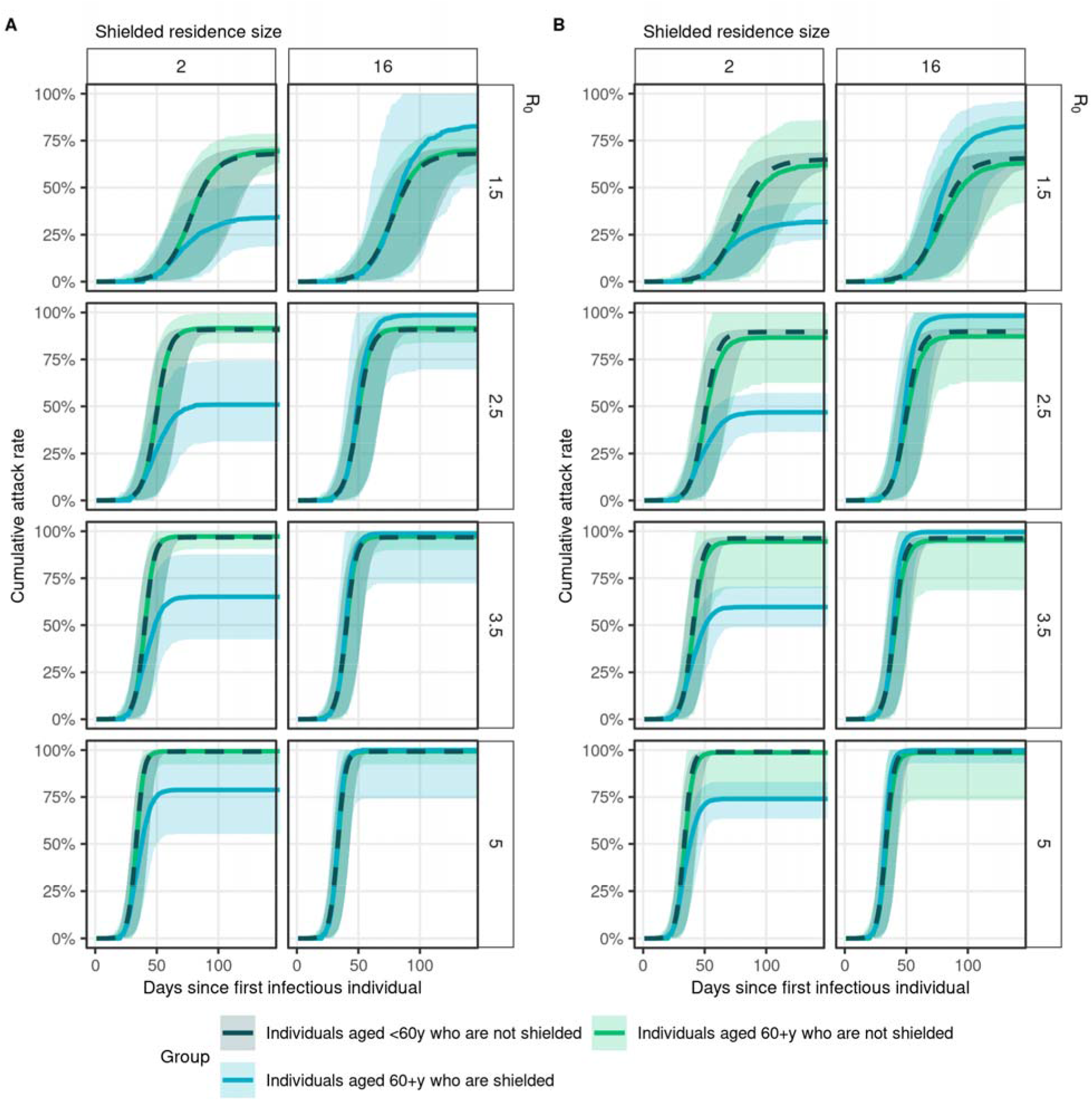
Cumulative infection risk by shielded status, stratified by shielded residence size (in facet columns) and *R*_0_ (in facet rows). Panel A shows scenarios where 20% of the high-risk population was Figure 2. Cumulative infection risk by shielded status, stratified by shielded residence size (in facet shielded, while panel B shows scenarios where 80% of the high-risk population was shielded. In each scenario, lines represent the median attack rate across 1000 model runs at each timepoint, whereas corresponding 95% uncertainty intervals are shown by shaded areas of the same colour. Estimates for (unshielded) low-risk individuals are shown in dark-green and a dashed line. Unshielded high-risk individuals are shown in light-green, while shielded high-risk individuals are shown in blue. In all scenarios shown, 0% of the population were immune at the start of the simulation, and contacts between shielded and unshielded individuals are reduced by 80%.

**Supplemental Figure 3.**
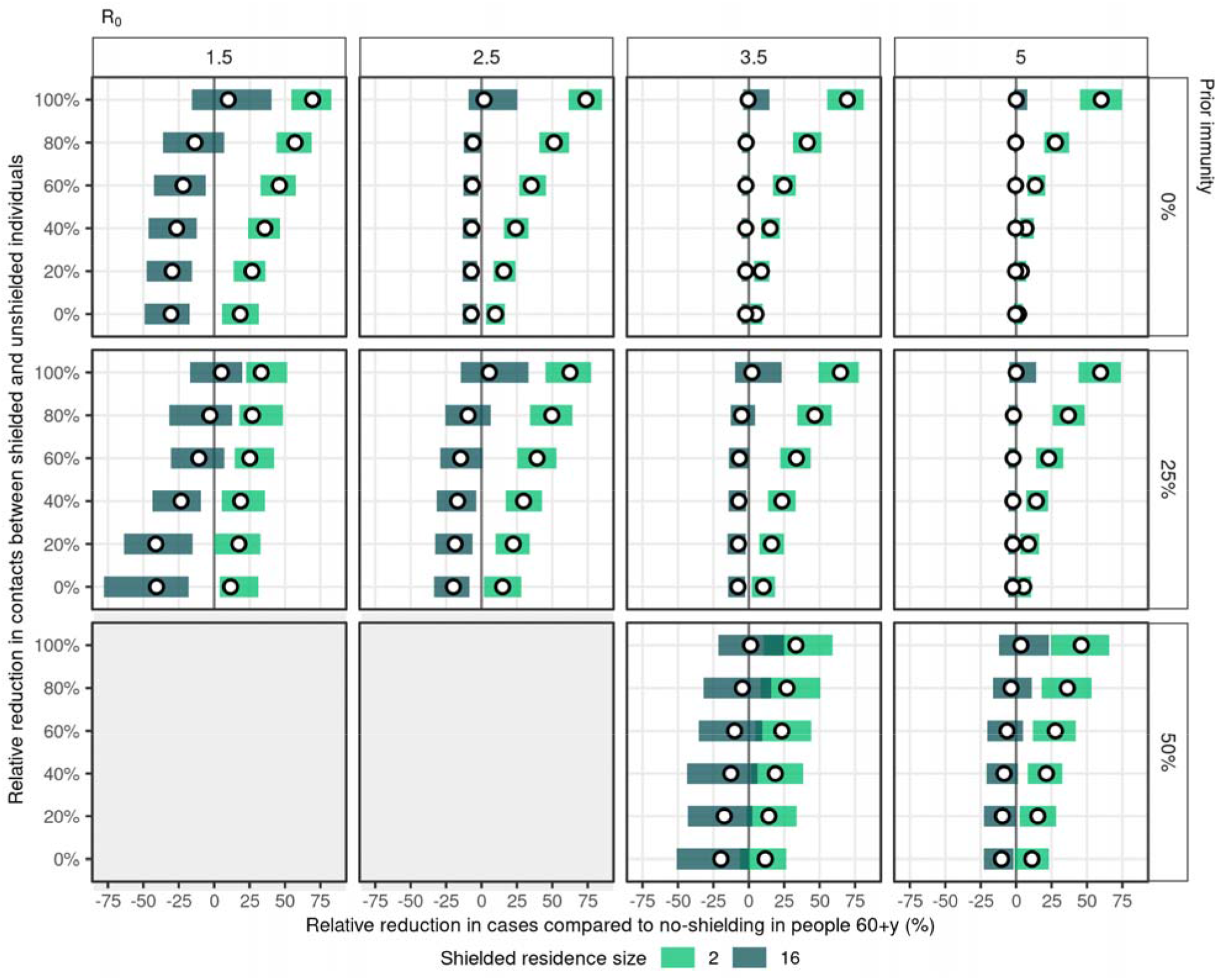
Relative reduction in cumulative infection risk in shielded high-risk individuals compared to no-shielding, by shielded residence size (shaded by colour), *R*_0_ (in facet columns), and the proportion immune at the start of the simulation (in facet rows). Points represent median estimates over 1000 model runs, while bars represent their 95% uncertainty intervals. In all scenarios shown, contacts between shielded individuals within the same shielded residence remain unchanged, symptomatic individuals who are shielded do not reduce their effective contacts, and no other mitigation measures are implemented. Contacts between shielded and unshielded individuals are reduced by the levels on the y-axis. Estimates to the left of the thick vertical grey line correspond to an increase in infection risk after shielding compared to no shielding. In all scenarios, 100% of high-risk individuals were shielded. There were not enough model iterations with large enough outbreaks to implement shielding in scenarios with an *R*_0_ of 1.5 or 2.5 and 50% prior immunity, so estimates are not available for these scenarios.

